# External validation of six clinical models for prediction of unknown chronic kidney disease in a German population

**DOI:** 10.1101/2021.12.23.21268085

**Authors:** Susanne Stolpe, Bernd Kowall, Denise Zwanziger, Mirjam Frank, Karl-Heinz Jöckel, Raimund Erbel, Andreas Stang

## Abstract

**Background:** Chronic kidney disease (CKD) is responsible for large personal health and societal burdens. Screening populations at higher risk for CKD is effective to initiate earlier treatment and decelerate disease progress. We externally validated clinical prediction models for unknown CKD that might be used in population screening.

**Methods:** We validated six risk models for prediction of unknown CKD using only non-invasive parameters. Validation data came from 4,185 participants of the German Heinz-Nixdorf-Recall study (HNR), drawn in 2000 from a general population aged 45-75 years. We estimated discrimination and calibration using the full model information, and calculated the diagnostic properties applying the published scoring algorithms of the models using various thresholds for the sum of scores.

**Results:** The risk models used four to nine parameters. Age and hypertension were included in all models. Five out of six c-values ranged from 0.71 to 0.73, indicating fair discrimination. Positive predictive values ranged from 15% to 19%, negative predictive values were >93% using score thresholds that resulted in values for sensitivity and specificity above 60%.

**Conclusions:** Most of the selected CKD prediction models show fair discrimination in a German general population. The estimated diagnostic properties indicate that the models are suitable for identifying persons at higher risk for unknown CKD without invasive procedures.

## Background

Prevalence of chronic kidney disease (CKD), defined by a chronically impaired renal function, is growing worldwide and a challenge for public health. In Germany, prevalence of a decreased renal function is up to 11.5 %^1^. Patients with CKD are at higher risk of cardiovascular comorbidities, hospitalization, end stage renal disease (ESRD) and premature death^2^. As a CKD cannot be cured, treatment aims at monitoring CKD risk factors – especially hypertension and blood glucose – to decelerate its progression and to prevent the incidence of secondary diseases^3^. Early diagnosis of a prevalent CKD can support these efforts. Despite the high prevalence and relevance for public health, public and patient awareness for CKD is low. The main reason for the low awareness is that CKD remains asymptomatic until reaching more serious stages. Moreover, a declining renal function is a physiological sign of older age, which often hinders physicians to designate an impaired renal function in older age as CKD.^4,5^

In Germany, fewer than 50% of patients with CKD -even with hypertension or cardiovascular disease - knew about their condition ^6,7^. In the USA, CKD unawareness is even more prevalent^8^. The high unawareness for CKD is astonishing, as regular monitoring of renal function should be mandatory for patients with hypertension, diabetes or of older age^3,9-11^. However, in general practice, even in patients with diabetes or hypertension, renal function, blood glucose or blood pressure are not regularly monitored ^12,13^ and risk factors are inadequately controlled ^14,15^.

According to Tonelli et al. most of the principles for population screening -formulated in a Delphi process among experts-fit to screening for CKD^16^. Screening for CKD has already been shown to be cost-effective in detecting unknown cases in population subgroups at higher risk for CKD ^17-19^. In a simulation study, it was shown that CKD prediction scores can be cost-effectively used to initially identify people at higher risk for incident CKD, and to screen these subsequently for CKD by testing for albuminuria ^20^. Prediction models suitable for identifying people at higher risk for CKD should be easy to apply, preferentially using non-invasive parameters only. Many CKD risk models of different complexity have already been developed – regarding the prediction of incidence, prevalence and progression to ESRD ^21^. However, missing external validation might frequently hinder the implementation into practice ^22,23^.

The aim of this study was, to externally validate prediction models, that estimate the probability of a prevalent unknown CKD using non-invasive parameters only in a German general population.

## Material and methods

### CKD prediction models

Starting with a review on CKD prediction models from 2012 ^21^, we searched the literature for further models that comprise only clinical information for estimating the risk of prevalent CKD. We identified five models that meet these criteria ^24-28^. Among these, Bang reported two versions of a model developed in the same population: SCORED and modified SCORED (Table 1). For these models, a scored version as self-completing questionnaire is published ^25^. The intercepts for the SCORED models and the models by Kwon and Thakkinstian had not been published. For the SCORED and the Kwon model, we were able to get the missing information about the intercept from the authors. For the Thakkinstian model we used the intercept published in a validation study ^23^. As the Kearns model estimated an unrealistic high risk for CKD in our validation population, we contacted the author and learned that the age-parameter should have had been centered (by subtracting the value 46.72) prior to the division of age by 10). This had not been reported in the manuscript. Further, no cutoffs or scoring rule for the Kearns-Model had been described, although sensitivity and specificity information had been estimated in the paper. As we could get no information on the applied rules for scoring the model parameters, we were not able to estimate the diagnostic properties of the Kearns model in our validation cohort.

**Table 1.** Identified prediction models for CKD: included parameters and their coefficients.

|  | Bang<br>SCORED | Bang „modified<br>SCORED“ | Kearns | Kshirsagar | Kwon | Thakkestian |
| --- | --- | --- | --- | --- | --- | --- |
| No of parameters | 9 | 7 | 5 | 8 | 7 | 4 |
| Intercept | -5.40<sup>a</sup> | -5.38<sup>a</sup> | -3.63 | -3.30 | -6.53<sup>a</sup> | -2.8<sup>b</sup> |
| Age (yrs) | 1.55 [50-59]<br>2.31 [60-69]<br>3.23 [$\geq 70$] | 1.55 [50-59]<br>2.29 [60-69]<br>3.29 [$\geq 70$] | 1.075 [Per 10 yrs<sup>c</sup>]<br>-0.01 [age<sup>2</sup>/10 yrs<sup>c</sup>]<br>0.104 [age < 50] | 0.63 [50-59]<br>1.33 [60-69]<br>1.46 [$\geq 70$] | 1.16 [50-59]<br>1.91 [60-69]<br>2.71 [$\geq 70$] | 0.6 [50-59]<br>1.4 [60-69]<br>2.1 [$\geq 70$] |
| Sex – Female | 0.29 | 0.34 | 0.73 | 0.13 | 0.4 |  |
| Anemia | 0.93 |  |  | 0.48 | 0.94 |  |
| Hypertension | 0.45 | 0.47 | 0.74<br>+ age <50: 0.56 | 0.55 | 0.48 | 0.8 |
| Diabetes | 0.44 | 0.47 |  | 0.33 | 0.73 | 0.9 |
| Ischemic heart disease or stroke (Hx)* | 0.59 | 0.67 |  | 0.26 | 0.60 |  |
| Heart failure (hx) | 0.45 | 0.51 | 0.86<br>CHF + age <50: 0.29 | 0.50 |  |  |
| Ischemic heart disease (Hx) |  |  | 0.51<br>+ age <50: 0.13 |  |  |  |
| Peripheral vascular disease (Hx) | 0.74 |  |  | 0.41 |  |  |
| Proteinuria | 0.83 | 0.88 |  |  | 0.48 |  |
| Kidney stones (Hx) |  |  |  |  |  | 1 |

a Intercept according to personal information by H. Bang.
b Intercept estimated using the prevalence of CKD in the validation population
c age per 10 yrs = (age -46.72) / 10yrs

The number of parameters used in the CKD prediction models ranged from four (Thakkinstian) to nine (SCORED) (Table 1). Age and hypertension were the only predictors used in all models. The Kearns model relies heavily on age, using age as interaction term with other parameters as well. History of kidney stones and history of ischemic heart disease were used only once.

### Validation population

We used the German Heinz Nixdorf Recall Study (HNR), a population based cohort, ^29^ for external validation. Baseline data from 4,814 participants drawn from the general population aged 45-75 years in 2000 were available. We included all participants with a valid measurement of serum creatinine (N=4,789).

### Measurement of variables

In HNR, all laboratory data had been analyzed centrally in the laboratory of the university hospital of Essen. Serum creatinine (according to Jaffé) was determined on a Siemens Healthcare Diagnostics ADVIA Chemistry. Serum creatinine was not standardized to isotope dilution mass spectrometry. Hypertension was defined as either a blood pressure of at least 140mmHg systolic or at least 90mmHg diastolic or taking antihypertensive medication. Blood pressure cut-offs were selected according to the cut-offs used to define hypertension in the validated risk models. Diabetes was defined according to the respective definitions used in the risk models: either self-reported prevalent diabetes ^25,26^ or using a combination of known diabetes or taking antidiabetic drugs ^27,28^. Albuminuria was defined as albumin/creatinine ratio (ACR) ≥30mg/dl. In all models except the Kwon model, anemia was coded if hemoglobin levels were <12 g/dl. In the Kwon model, the threshold for hemoglobin was <12 g/dl for women and <13 g/dl for men. Peripheral vascular artery disease was defined according to clinical information.

### Definition of chronic kidney disease in development and validation populations

In all validated risk models, CKD was defined as an estimated glomerular filtration rate (eGFR) <60ml/min/1.73m^2^ calculated by the Modification of Diet in Renal Disease (MDRD) equation.

All models had been developed to estimate the risk of unknown prevalent CKD stages 3 or more. Therefore, we defined CKD as eGFR <60ml/min/1.73m^2^ accordingly. We used the CKD-Epi equation for calculating eGFR, as recommended by the Kidney Disease Improving Global Outcomes (KDIGO) ^30^. In Germany, the CKD-Epi equation is widely used to report eGFR with laboratory results. As sensitivity analysis, we calculated the eGFR with the MDRD and the new Full-Age-Spectrum (FAS) equation (equations listed in supplement S2+S3) ^31^. We used the respective creatinine-based equations, because cystatin c measurements are not widely available in general practice.

### Handling of missing values

We did a complete case analysis regarding all predictors used in the identified models, leaving 4,185 participants in HNR. The Thakkinstian score was validated in a subsample of HNR with information on the parameter ‘history of kidney stones’ that is used in this model only (N=3,433).

### Statistical analysis

The models’ discrimination was estimated by the c-value and the Tjur coefficient ^32^. The Tjur coefficient is the difference between the mean predicted probability in cases and in non-cases. The higher this difference the better the discriminative ability of a score. Calibration was assessed graphically.

As measures for overall performance, we estimated the mean average prediction error (MAPE) and the Scaled Brier Score ^23,32^. MAPE averages the deviations between the prediction (ranging between 0 and 1) and the respective true value of zero or 1. The smaller the MAPE, the better the prediction. The Scaled Brier Score is calculated by the squared difference between the prediction and the true value of outcome (=Brier Score) divided by the product of the mean prediction value and 1-mean prediction value ^33^. It ranges from 0 to 1 representing 0% to 100% and is similarly interpreted as Pearson’s R^2^, indicating the rate of variability explained by the model.

We estimated sensitivity, specificity and predictive values of the models after scoring the model parameters using the cut-offs reported by the authors. The optimal threshold for the validation population was identified with the Youden index.

Additionally, we calculated the rate of expected to observed cases (E/O-proportion) for the thresholds used. An E/O-proportion close to 1 indicates agreement between the number of expected cases according to the models’ cut-offs and observed cases, an E/O-proportion >1 indicates overestimation of CKD risk.

All statistical calculations were done using SAS 9.4.

## Results

### Validation of identified risk models in the German HNR study

Compared to the development populations of the validated risk models, the participants in the HNR study were older (mean age 59.6 compared to 44.2 to 57 years), and reported hypertension more often (59% compared to 15 to 36%) (Table 3). Prevalence of CKD in HNR calculated by the MDRD-equation was 8.6%. It was higher than in the development populations of the Bang (SCORED) (5.4%), Kearns (6.8%) and Kwon (4.6%) models. In the US based population used for deriving the Kshirsagar Score, CKD prevalence was 16.9%. Thakkinstian reported a prevalence of 17.5%, however, in contrast to the other studies, CKD here comprises CKD stages 1-5.

**Table 2.** Characteristics of the validation and development populations of the CKD risk models (means and standard deviation (SD) or percent (%)).

|  | Heinz-Nixdorf-Recall-Study | Development populations for identified CKD prediction models |  |  |  |  |
| --- | --- | --- | --- | --- | --- | --- |
|  |  | Bang (SCORED, modified<br>SCORED) 2007 | Kearns 2013 | Kshirsagar 2008 | Kwon 2011 | Thakkestian 2011 |
| Country | Germany | USA, 72% white | UK, 30% white,<br>53% unknown | USA, 78% white | Korea | Thailand |
| Size of population (N) | 4,185 | 8,530 | 743,935 | 9,470 | 6,565 | 3,459 |
| CKD defined by | GFR<60 | GFR<60 | GFR<60 | GFR<60 | GFR<60 | KDIGO CKD1-5 |
| Prevalence of CKD | 8.6% (MDRD)<br>9.2% (CKD-Epi) | 5.4% (MDRD) | 6.8% (MDRD) | 16.9% (MDRD) | 4.6% (MDRD) | 17.5 (CKD1-5,<br>MDRD) |
| Handling of missings | Excluded | Excluded | Missing BP:<br>no hypertension | Not reported | Excluded | Not reported |
| Female | 50.5% | 52% | 50% | 56% | 50% | 54.5% |
| Age, mean (SD) | 59.6 (7.8) | 46.0 (31.4) | 46.7 (18.2) | 57 (9) | 44.2 (32.4) | 45.2 (46.5) |
| Age, range | 45-75 | 20-85 | ≥18 | 45-64 | ≥19 | ≥18 |
| BMI, mean (SD) | 27.8 (4.6) | 28.0 (12.9) | - | 27 (5) | 23.6 (8.1) | 24.0 (11.8) |
| Smoking | 23.3% | 20% | 20% | 10% | 27.5<sup>a</sup> | 35.9% |
| Diabetes | 7.9% | 8% | 5% | 9% | 8.3%<sup>b</sup> | 11.9% |
| Hypertension | 59.2% | 34% | 15% | 36% | 22.5% | 27.5% |
| Ischemic heart disease<br>or stroke | 6.9% | 4.9% | 2% stroke, 4% IHD | 8% | 3.2% (IHD or<br>stroke) | 3.4% (heart disease) |
| Heart failure | 3.5% | 2.1% | 1% | 0.7% | - | - |
| Proteinuria | 1.7% | 10% | - | - | 10.3% | - |
| PVD | 2.3% | 2.7% | 1% | 4% | - | - |
| Kidney stones | 12.0%<br>(N=3,398) | - | - | - | - | 5.0% |
| Serum creatinine<br>mean (SD) | 0.93 (0.2) | 0.89 (0.4) | - | 0.8 (0.2) | 0.9 (8.1) | 1.1 (1.18) male,<br>0.8 (1.18) female |
| eGFR, mean (SD) | 79.1 (17.6)<br>(CKD-Epi) | 94.0 (48.9)<br>(MDRD) | - | - | 85.9 (56.7)<br>(MDRD) | - |
| Anemia | 2.1% (Hb <12) | 2.7%<sup>c</sup> | - | - | 8.1% | - |

BP: blood pressure; CKD: chronic kidney disease; CVD: cardiovascular disease; IHD: ischemic heart disease; KDIGO: Kidney Disease Improving Global Outcomes; MDRD: modification of Diet in Renal Disease; PVD: peripheral vascular disease
a: smoking defined as: >5 packs life time or current.
b: diabetes defined as: glucose ≥126 or anti-diabetic medication or insulin therapy.
c: anemia defined as: treatment of anemia.
d: history of bypass operation used as surrogate for heart failure.

**Table 3.**
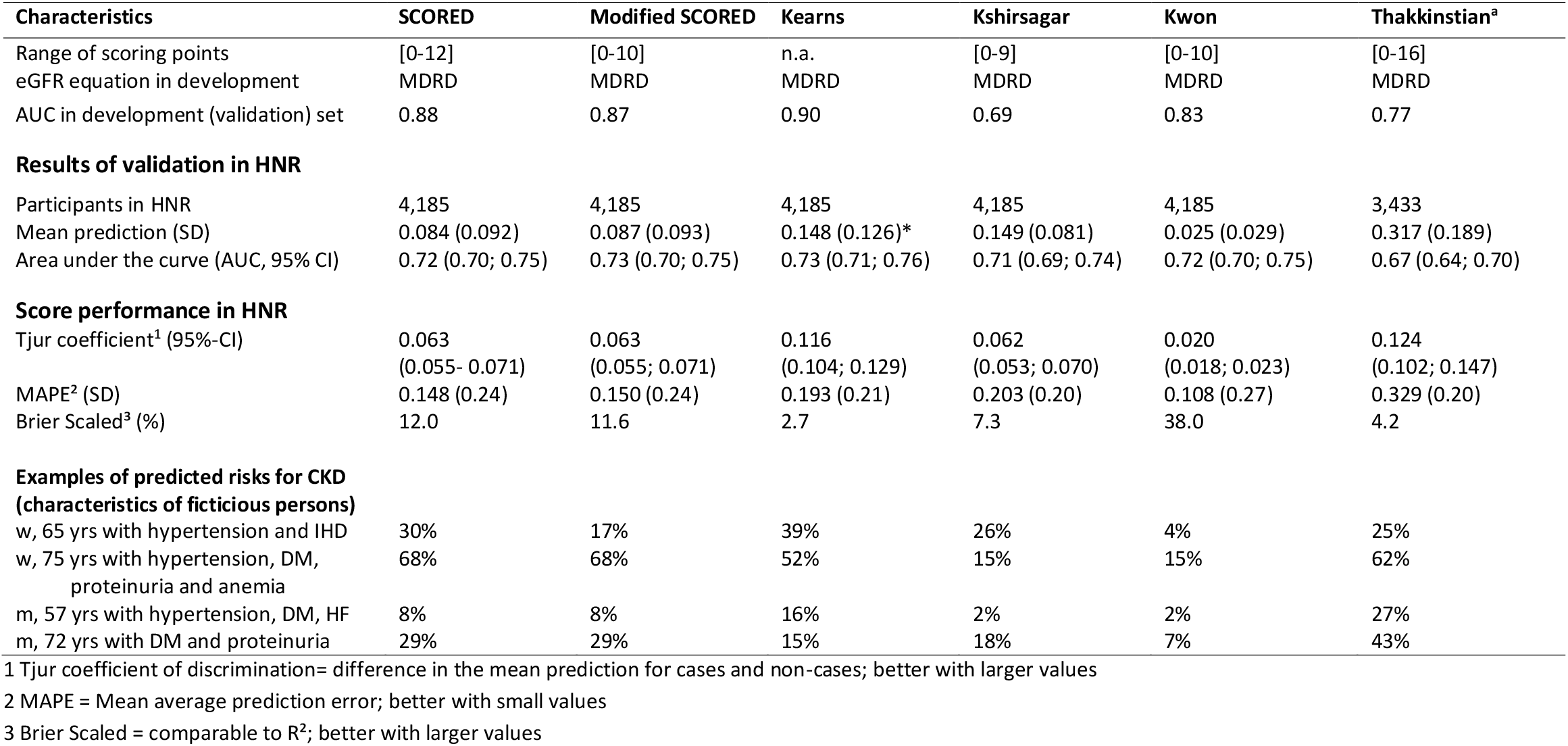
External validation of identified models to predict unknown CKD; validation data set: Heinz Nixdorf Recall study (Germany). CKD is defined as eGFR <60ml/min/1.73m^2^ using CKD-Epi equation for calculating eGFR. Measures presented with standard deviation (SD) or 95% confidence intervals (CI).

The mean estimated probability of prevalent CKD differed strongly and ranged from p=0.025 (Kwon) to 0.317 (Thakkinstian) (Table 3).

### Discrimination

Discrimination of all models but in the Kshirsagar model was lower in HNR compared to the development data sets (Table 3). C-values ranged from 0.67 (Thakkinstian) to 0.73 (modified SCORED) using CKD-EPI equation for defining CKD. With MDRD equation, c-values were <0.7 for all scores. The FAS equation yielded a better discrimination (c-values 0.74-0.80) (see supplement table S4). The Tjur-coefficient was largest and indicated best discrimination of prediction for the Thakkinstian score (0.124) and the Kearns model (0.116).

### Calibration

The calibration plots showed reasonable fit only with the Bang models (SCORED, modified SCORED). CKD risk was overestimated in persons with lower CKD risk and overestimated in persons with higher risk. Risk of unknown CKD was generally overestimated by the Kearns, Kshirsagar and Thakkinstian model. The Kwon model underestimated the probability of unknown CKD. Calibration to the HNR population was poor in all but the SCORED models (see supplementary file).

The Kwon score yielded the smallest mean average prediction error with 0.108 compared to the Thakkinstian score resulting in the largest MAPE with 0.329. The Brier Scaled which can be interpreted as R^2^ indicated that the Kwon score prediction fitted best to the population.

Applying the published scoring rules for the models to the HNR study, the E/O-proportion depended strongly on the eGFR equation used. For example, using a threshold of six points, the ‘modified SCORED’ had an E/O-proportion of 1.17 (95%-CI: 1.06; 1.28) with the CKD-Epi equation, 1.55 (1.42; 1.68) with the MDRD and 0.91 (0.84; 0.99) with the FAS equation (supplement table 4, S4).

**Tab. 4.**
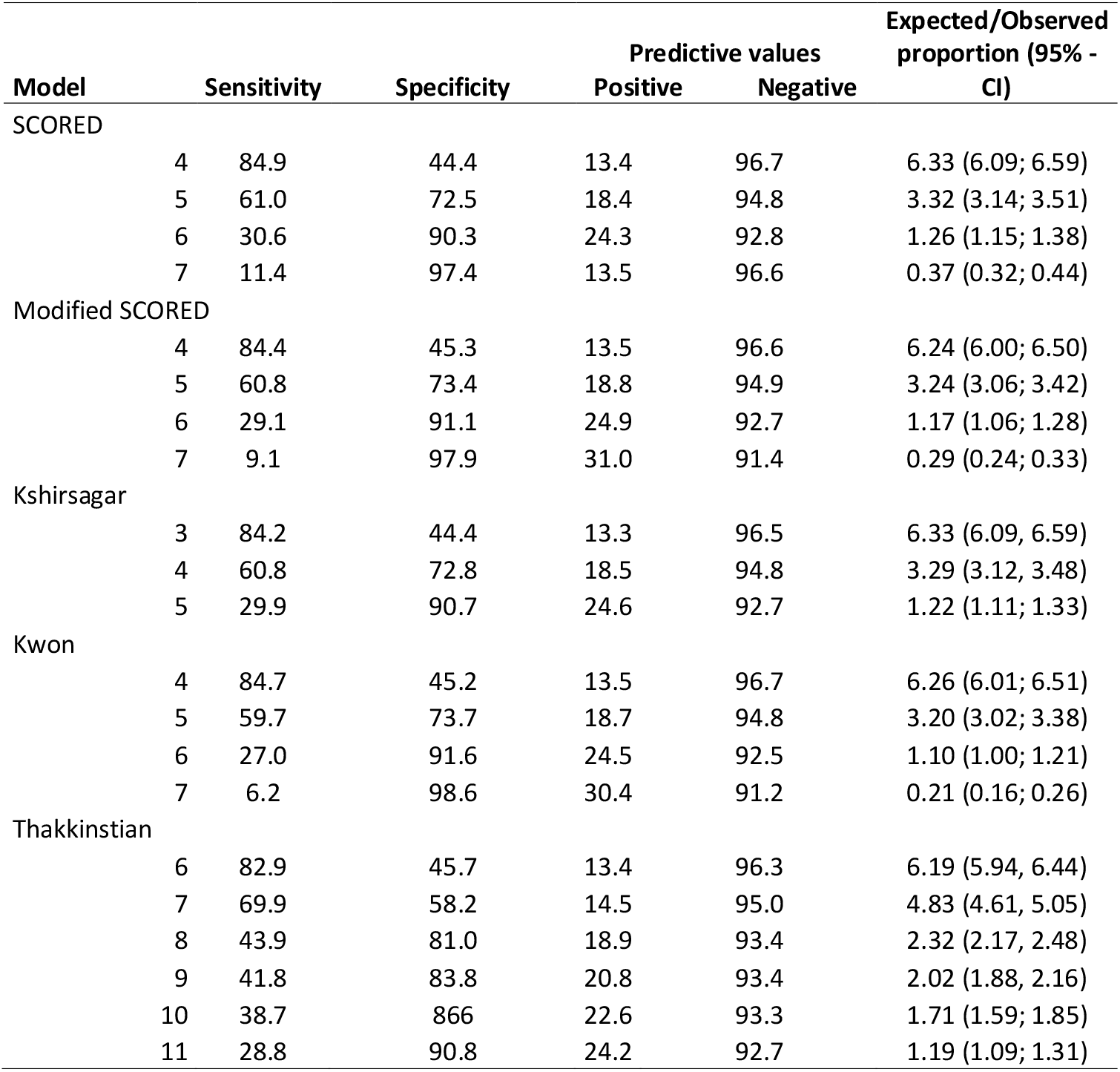
Diagnostic criteria of validated models for various thresholds (sensitivity, specificity and predictive values as well as proportions of expected to observed cases with 95%-confidence intervals) for CKD. CKD defined as eGFR <60m/min/1.73m^2^ calculated with CKD-Epi equation; CKD prevalence 9.2%.

### Diagnostic criteria

Thresholds of the scored models that resulted in a sensitivity >60% and specificity >60% (five points for the Bang and the Kwon models, four points for the Kshirsagar model and 7 points for the Thakkinstian model) resulted in positive predictive values (PPV) between 15% and 19%. Negative predictive values (NPV) were similar for all models independent of a chosen threshold and ranged between 93% and 95% (Tab. 4). Using the FAS-equation yielded the highest sensitivity and specificity for all thresholds compared to the CKD-EPI or MDRD equation (see supplement table S3a/b).

## Discussion

We externally validated six prediction models which estimate the probability for prevalent unknown CKD without any laboratory measurement. Most models yielded c-values of about 0.72 in the German HNR validation cohort. The calibration to the validation data set was reasonable only for the Bang (SCORED) models. A PPV of about 19% in a general population as estimated with all models but the Thakkinstian model indicates a good suitability as tools to identify patients at higher risk for whom further CKD diagnostic would be advisable.

### Current use of risk models in CKD

Only few existing CKD prediction models are in use, such as the Kidney-Failure-Risk-Equation for prediction of renal failure. ^34,35^. To prevent progression to serious consequences of CKD such as ESRD or cardiovascular diseases, early diagnosis of CKD is necessary. Due to the asymptomatic progress of the disease even patients in later stages of CKD are often undiagnosed and therefore untreated ^8^

Contrasting to screening for CKD in the general population, case finding in populations at higher risk has been proven cost-effective ^36,37^. Existing CKD risk models have not yet been applied in programs aiming at identifying persons at higher risk for CKD. ^18,38^. The Kshirsagar and the SCORED model were used in a simulation study that proved cost-effectiveness in identifying persons for screening for early stages of CKD ^20^, SCORED was evaluated as screening tool for CKD in a small number of participants (N=172) as alternative to regular CKD screening protocols ^39^.

Missing external validation of published CKD risk models can hinder clinical implementation. The SCORED, the Kwon-, Khirsagar- and the Thakkinstian-model have already been externally validated in 2016 in a UK population, but not with regard to the probability of prevalent CKD ^22^. Also, models using parameters that are either unfamiliar, unusually scaled, complicated to calculate or costly to collect (e.g. genetic information) ^40^ or that do not reflect the general opinion in clinic about risk factors have a low probability of being used in clinical routine. ^41,42^. Most of the identified models comprise familiar predictors which reflect the current knowledge about CKD risk factors. Only the Kearns and the Thakkinstian model do not use diabetes or sex for CKD prediction– parameters which usually would be regarded as relevant for estimating the risk for CKD.

### Validation results

Although the number and type of parameters used in the identified models differed, all but the Thakkinstian model showed fair discriminative properties in the HNR cohort with c-values ranging from 0.72 – 0.74. These c-values can be judged as satisfactory regarding the non-invasive and dichotomous nature of their predictors that facilitate potential implementations. Taking the small age range of the HNR cohort compared to the development populations of most of the validated scores into account, c-values could have been expected to be lower than in validation populations with full-age-spectrum ^43^ as, within a small age-range, it is more difficult to discriminate cases and non-cases when age is the most relevant prediction factor.

Calibration plots revealed a slight underestimation of CKD risk for the two Bang models. For all other models, calibration was poor. We recommend a re-calibration of the intercept ^44^ if the estimated probabilities are of interest in regard to a clinical implementation of a model. In our estimation of the diagnostic properties we relied on the sum of scoring points for the parameters.

### Potential implementation of the risk models

As the selected models do not use any laboratory or genetic information to estimate the risk for a prevalent CKD, these models can be used in screening scenarios where laboratory or genetic information would be too difficult or too expensive to get. Using the scoring rules for the models regarding the answers to the model parameters would enable to implement these models as self-completing check-list tool for patients which can easily be evaluated in a screening scenario^43^. However, acceptance of a prediction model is dependent on its face validity which means that the model parameters describe known risk factors. The validity of the Thakkinstian model without consideration of sex or of the Kearns model which does not imply diabetes might be questioned by physicians. On the other hand, proteinuria as a known risk factor is included in the SCORED and the Kwon model, but whether patients know for sure whether they had blood in their urine can be doubted.

Nevertheless, in a German general population, the modified SCORED and the Kwon model had good external validity and diagnostic properties. We think, that both models are suitable to identify people at higher risk for CKD at low cost if implemented as web based tool or distributed as paper questionnaire on information leaflets for example in public places or at health institutions. People who learn that they have a higher risk for CKD according their answers to the questionnaire may inform their GP who can decide to initiate further CKD diagnostic. We think this pragmatic approach can contribute to higher awareness for CKD, leading to earlier diagnosis and treatment.

### Strengths

The HNR cohort is of high data quality and has been the base for many publications so far. All relevant parameters of the models have been available for the external validation. To our knowledge we are the first to externally validate prediction models for unknown CKD.

### Limitations

We did not intend to do a systematic review on all CKD models suitable for risk estimation for unknown CKD. Therefore it might be possible that we did not include all existing model. However, to our knowledge, we were the first to evaluate all the selected models in regard to their ability to predict prevalent CKD. We think that we herewith support potential implementations of these models.

The MDRD equation used in our sensitivity analyses has its weakness in a limited validity with eGFR levels >60ml/min/1.73m^2^. This however does not affect our validation results, as all models estimate the risk for CKD stage 3 or more which is defined by lower eGFR levels.

## Conclusions

External validation of risk models for unknown CKD yielded fair discrimination in a German population-based cohort. Calibration to the data was satisfactorily only for some scores. Diagnostic properties show that the models can be useful in screening scenarios to identify people at higher risk for CKD. As only non-invasive parameters are used, they can easily be implemented as tool for patient self-assessment of CKD risk.

## Data Availability

Due to data security reasons (ie, data contain potentially participant identifying information), the HNR Study does not allow sharing data as a public use file. Data requests can be addressed to.

## Ethics approval and consent to participate

The Heinz-Nixdorf-Study has been approved by the ethic committee of the university hospital Essen on 12. May 1999. All participants gave their written informed consent before participating in the studies.

## Participant protection

Data is anonymized according to the data protection act, individual participants are not identifiable.

## Authors’ contributions

Research idea and study design: SS, AS; data acquisition: KJ, MF, DZ, RE; analysis/interpretation: SS, BK; statistical analysis: SS; supervision or mentorship: AS, BK. Each author contributed important intellectual content during manuscript drafting or revision and agrees to be personally accountable for the individual’s own contributions and to ensure that questions pertaining to the accuracy or integrity of any portion of the work, even one in which the author was not directly involved, are appropriately investigated and resolved, including with documentation in the literature if appropriate.

## Acknowledgement

The authors express their gratitude to all study participants of the Heinz Nixdorf Recall (HNR) Study, the personnel of the HNR study center and the EBT-scanner facilities, the investigative group and all former employees of the HNR study. The authors also thank the Advisory Board of the HNR Study: T. Meinertz, Hamburg, Germany (Chair); C. Bode, Freiburg, Germany; P.J. de Feyter, Rotterdam, Netherlands; B. Güntert, Hall i.T., Austria; F. Gutzwiller, Bern, Switzerland; H. Heinen, Bonn, Germany; O. Hess (†), Bern, Switzerland; B. Klein (†), Essen, Germany; H. Löwel, Neuherberg, Germany; M. Reiser, Munich, Germany; G. Schmidt (†), Essen, Germany; M. Schwaiger, Munich, Germany; C. Steinmüller, Bonn, Germany; T. Theorell, Stockholm, Sweden; and S.N Willich, Berlin, Germany.

## Supporting information

Supporting file includes:

S1: Equations for eGFR estimation used in the analyses

S2: Equation of prediction models externally validated

S3: Fig S1: Calibration of the six models using CKD-Epi equation to define CKD 3-5

S4: Performance indicators for selected CKD risk models; CKD defined as eGFR <60ml/min/1.73m^2^ calculated with FAS and MDRD equation

S5: Diagnostic criteria for prediction models for various threshold for CKD defined by MDRD and FAS equations ((sensitivity, specificity and predictive values and the respective proportions of expected to observed cases and 95%-confidence intervals).

