## supplement for "External validation of six clinical models for prediction of unknown chronic kidney disease in a German population"

**Contents:**

S1: Equations for eGFR estimation used in the analyses

S2: Equation of prediction models externally validated

S3: Fig S1: Calibration of the six models using CKD-Epi equation to define CKD 3-5

S4: Performance indicators for selected CKD risk models; CKD defined as eGFR <60ml/min/1.73m² calculated with FAS and MDRD equation

S5: Diagnostic criteria for prediction models for various threshold for CKD defined by MDRD and FAS equations ((sensitivity, specificity and predictive values and the respective proportions of expected to observed cases and 95%-confidence intervals).

**S1: Equations for eGFR estimation used in the analyses:**

SCr = serum creatinine

MDRD: 186 x (SCr) $^{-1..154}$ x age $^{-0.203}$ x 0.742 (if female) x 1.21 (if black)

CKD -EPI: 141 x min(SCr/k,1)$^{a}$ x max(SCr/k,1) $^{-1.209}$ x 0.993$^{age}$ [x 1.018 if female] [x 1,159 if black]

where k =0.7 for females and 0.9 for males, a= -0.329 for females and -0.411 for males

FAS: 107.3 /SCr/Qvalue [x 0.988 $^{(age-40)}$ if age>40 years],

where QValue is 0.85 for males aged 18 years, 0.88 for females aged 19 years , 0.90 for males aged ≥20 years, 0.69 for females aged 18 years and 0.70 for females aged ≥19 years.

**S2: Equation of prediction models externally validated:**

(CKD= chronic kidney disease, CVD = cardiovascular disease i.e. ischemic heart disease or stroke; PVD = peripheral vascular disease)

Predictors in () take 1 for event a and 0 otherwise.

SCORED (Bang 2007)

Probability (CKD) = 1/[1 + exp(-ß' * X)], where ß' * x = -5.4 + 1.55 * (age of 50-59 years) + 2.31 * (age of 60-69 years) + 3.23 * (age >=70 years) + 0.29 * (female) + 0.93 * (anemia) + 0.45 * (hypertension) + 0.44 * (diabetes) + 0.59 *(history of CVD) + 0.45 *(history of heart failure) + 0.74 * (PVD) + 0.83 * (proteinuria).

Modified SCORED

Probability (CKD) = 1/[1 + exp(-ß' * X)], where ß' * x = -5.38 + 1.55 * (age of 50-59 years) + 2.29 * (age of 60-69 years) + 3.29 * (age >=70 years) + 0.34 * (female) + 0.47 * (hypertension) + 0.47 * (diabetes) + 0.67 *(history of CVD) + 0.51 *(history of heart failure) + 0.88 * (proteinuria).

Kearns

Probability (CKD) = 1/(1+exp(-ß' * X)], where ß' * x = -3.63 + 1.075*(age-46.72)/10 -0.01*( age-46.72)/10 * age-46.72)/10)-0.734*male + 0.104*age_under_50 + 0.863*(history of heart failure) + 0.29*(history of heart failure and age_under50) + 0.74*hypertension + 0.56*(hypertension and age under50) + 0.51*(history of ischemic heart disease) + 0.13*(history of ischemic heart disease and age_under50))

Khirsagar

Probability (CKD) = 1/[1 + exp(-ß' * X)], where ß' * x = -3.30 + 0.63 * (age of 50-59 years) + 1.33 * (age of 60-69 years) + 1.46 * (age >=70 years) + 0.13 * (female) + 0.48 * (anemia) + 0.55 * (hypertension) + 0.33 * (diabetes) + 0.26 * (History of CVD) + 0.50 *(history of heart failure) + 0.41 * (PVD).

Kwon

Probability (CKD) = 1/[1 + exp(-ß' * X)], where ß' * x = -6.53 + 1.16 * (age of 50-59 years) + 1.91 * (age of 60-69 years) + 2.71 * (age >=70 years) + 0.40 * (female) + 0.94 * (anemia) + 0.48 * (hypertension) + 0.73 * (diabetes) + 0.60 * (History of CVD) + 0.48 *(proteinuria).

Thakkinstian

Probability (CKD) = 1/[1 + exp(-ß' * X)], where ß' * x = -2.80 + 0.6 * (age of 50-59 years) + 1.4 * (age of 60-69 years) + 2.1 * (age >=70 years) + 0.8 * (hypertension) + 0.9 * (diabetes) + 1 * (History of kidney stones).

**Fig. S1. Calibration plots prediction models for unknown CKD in the German Heinz-Nixdorf-Recall study (N=4.185).**

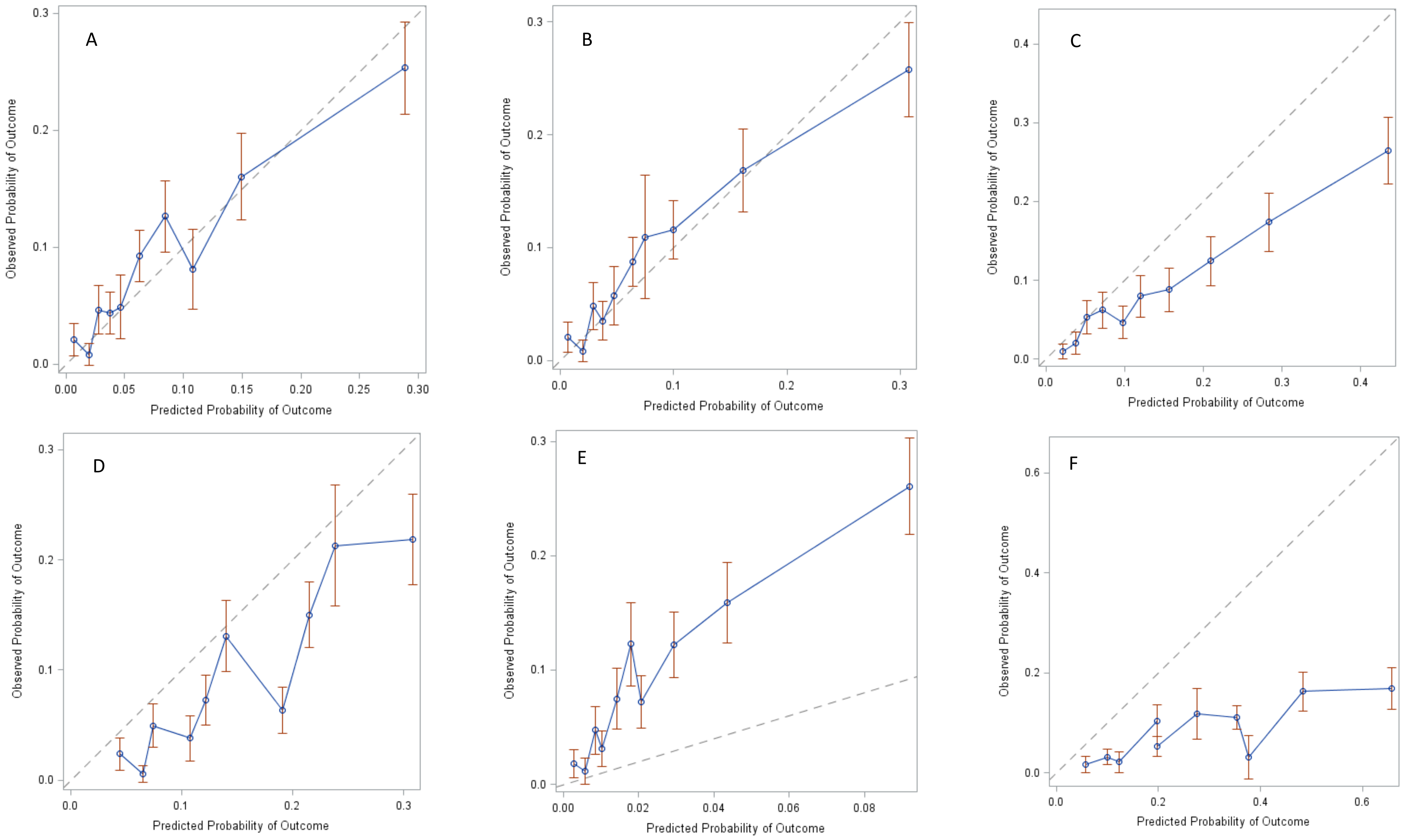


A: Bang SCORED (Bang et al. 2007), B: Bang Modified SCORED (Bang et al. 2007), C: Kearns Model (Kearns et al. 2013), D: Kshirsagar Model (Kshirsagar 2008), E: Kwon Model (Kwon et al. 2012), F: Thakkinstian Model (Thakkinstian et al. 2011).

**Table S4: Performance indicators for selected CKD risk models; CKD defined as eGFR <60ml/min/1.73m² calculated with FAS and MDRD equation**

MDRD:

| Characteristics | eGFR equation | **SCORED** | **Modified SCORED** | **Kearns** | **Kshirsagar** | **Kwon** | **Thakkinstian** | |
|---|---|---|---|---|---|---|---|---|
| Area under the curve (AUC) | | | | | | | | |
| CKD definition | MDRD | 0.68 (0.66; 0.71) | 0.69 (0.66; 0.71) | 0.70 (0.67; 0.72) | 0.68 (0.65; 0.71) | 0.69 (0.66; 0.71) | 0.63 (0.60; 0.67) | |
| | FAS | 0.79 (0.78; 0.81) | 0.80 (0.78; 0.81) | 0.80 (0.79; 0.82) | 0.78 (0.76; 0.79) | 0.79 (0.77; 0.81) | 0.73 (0.71; 0.75) | |
| Tjur-Coefficient (95%-CIl) | | | | | | | | |
| CKD-definition | MDRD | 0.053 (0.045; 0.061) | 0.053 (0.44; 0.60) | -- | 0.052 (0.043; 0.061) | 0.018 (0.015; 0.020) | 0.093 (0.071; 0.115) | 0.099 (0.074; 0.123) |
| | FAS | 0.083 (0.077; 0.089 | 0.084 (0.075; 0.092) | -- | 0.078 (0.071; 0.085) | 0.026 (0.024: 0.028) | 0.157 (0.140; 0.174) | 0.167 (0.149; 0.185) |

Modification in Diet in Renal Disease; FAS= For all age spectrum

**Tab. S5. Diagnostic criteria for prediction models for various threshold for CKD defined by MDRD and FAS equations (sensitivity, specificity and predictive values and the respective proportions of expected to observed cases and 95%-confidence intervals).**

| | FAS ( CKD prevalence 14.7%) | | | | | MDRD (CKD prevalence 8.6%) | | | | |
|---|---|---|---|---|---|---|---|---|---|---|
| | | | Predictive values | | | | | Predictive values | | |
| | Sensitivity | Specificity | Positive | Negative | Proportion expected/observed | Sensitivity | Specificity | Positive | Negative | Proportion expected/observed |
| **SCORED** | | | | | | | | | | |
| **4** | 89.9 | 47.2 | 22.6 | 93.3 | 4.14 (3.98; 4.30) | 81.4 | 43.9 | 12.0 | 96.2 | 7.04 (6.77; 7.32) |
| **5** | 68.0 | 75.9 | 32.6 | 93.3 | 2.37 (2.25; 2.49) | 56.4 | 71.9 | 15.9 | 94.6 | 4.03 (3.83; 4.24) |
| **6** | 36.4 | 92.6 | 45.9 | 89.5 | 0.97 (0.90; 1.05) | 28.3 | 90.0 | 21.0 | 93.0 | 1.65 81.53; 1.79) |
| **7** | 13.2 | 98.3 | 56.6 | 86.8 | 0.33 (0.29; 0.38) | 11.1 | 97.3 | 28.0 | 92.1 | 0.56 (0.49; 0.61) |
| **Modified SCORED** | | | | | | | | | | |
| **4** | 89.6 | 48.1 | 22.8 | 96.4 | 4.08 (3.93; 4.25) | 80.8 | 44.8 | 12.1 | 96.1 | 6.96 (6.69; 7.23) |
| **5** | 67.9 | 76.8 | 33.4 | 93.3 | 2.32 (2.20; 2.44) | 56.1 | 72.7 | 16.2 | 94.6 | 3.94 (3.74; 4.16) |
| **6** | 33.9 | 93.3 | 46.3 | 89.2 | 0.91 (0.84; 0.99) | 26.9 | 90.8 | 21.6 | 93.0 | 1.55 (1.42; 1.68) |
| **7** | 10.9 | 98.7 | 59.3 | 86.6 | 0.29 (0.25; 0.33) | 8.6 | 97.9 | 27.4 | 91.9 | 0.49 (0.42; 0.57) |
| **Kshirsagar** | | | | | | | | | | |
| **3** | 89.4 | 47 | 22.5 | 96.3 | 3.98 (3.82, 4.14) | 80.8 | 43.9 | 11.9 | 96.1 | 6.77 (6.51, 7.05) |
| **4** | 67.7 | 76.1 | 32.7 | 93.2 | 2.07 (1.96, 2.19) | 56.1 | 72.1 | 15.9 | 94.6 | 3.52 (3.33, 3.72) |
| **5** | 35.1 | 92.9 | 45.9 | 89.3 | 0.76 (0.70, 0.84) | 27.5 | 90.4 | 21.2 | 93 | 1.30 (1.19, 1.42) |
| **Kwon** | | | | | | | | | | |
| **4** | 89.6 | 47.9 | 22.8 | 96.4 | 4.10 (3.94; 4.26) | 81.4 | 44.7 | 12.2 | 96.2 | 6.98 (6.71; 7.26) |
| **5** | 67.0 | 77.0 | 33.4 | 93.2 | 2.29 (2.18; 2.42) | 55.3 | 73.0 | 16.2 | 94.5 | 3.91 (3.71; 3.85) |
| **6** | 33.0 | 93.8 | 47.6 | 89.1 | 0.88 (0.80; 0.95) | 25.3 | 91.3 | 21.5 | 92.8 | 1.49 (1.37; 1.62) |
| **7** | 8.0 | 99.2 | 62.0 | 86.3 | 0.24 (0.21; 0.29) | 6.1 | 98.5 | 27.8 | 91.8 | 0.42 (0.36; 0.49) |
| **Thakkinstian** | | | | | | | | | | |
| **6** | 89.6 | 48.7 | 23.1 | 96.5 | 3.98 (3.82, 4.14) | 77.2 | 45.0 | 11.7 | 95.5 | 6.62 (6.36, 6.89) |
| **7** | 77.3 | 61.2 | 25.5 | 94.0 | 2.07 (1.96, 2.19) | 64.2 | 57.4 | 12.4 | 94.5 | 5.16 (4.93, 5.40) |
| **8** | 50.7 | 83.7 | 34.8 | 90.8 | 0.76 (0.70, 0.84) | 38.1 | 80.2 | 15.3 | 93.2 | 2.48 (2.32, 2.65) |

| | | | | | | | | | | |
|---|---|---|---|---|---|---|---|---|---|---|
| **9** | 48.1 | 86.5 | 38.0 | 90.7 | 3.98 (3.82, 4.14) | 35.8 | 83.1 | 16.6 | 93.2 | 2.16 (2.01, 2.31) |
| **10** | 45 | 89.3 | 41.8 | 90.4 | 2.07 (1.96, 2.19) | 32.8 | 85.8 | 17.9 | 93.1 | 1.83 (1.70, 1.98) |
| **11** | 34.8 | 93.1 | 46.4 | 89.3 | 0.76 (0.70, 0.84) | 24.2 | 90.3 | 19.0 | 92.7 | 1.28 (1.16, 1.40) |
